# Transferability of European-derived cardiometabolic polygenic risk scores in the South Asians and their interplay with family history

**DOI:** 10.1101/2023.03.20.23287470

**Authors:** Emadeldin Hassanin, Carlo Maj, Peter Krawitz, Patrick May, Dheeraj Reddy Bobbili

**Affiliations:** Luxembourg Centre for Systems Biomedicine, University of Luxembourg, Esch-Sur-Alzette, Luxembourg; Institute for Genomic Statistics and Bioinformatics, University of Bonn, Bonn, Germany; Centre for Human Genetics, University of Marburg, Marburg, Germany; Wellytics Technologies Pvt Ltd, Hyderabad, India

**Keywords:** Type 2 diabetes, Family History, South Asians, Polygenic Risk, Coronary artery disease

## Abstract

**Background & Aims:** We aimed to investigate the effect of polygenic risk scores (PRSs) derived from individuals of European (EUR) ancestry on common diseases among individuals of South Asian (SAS) ancestry in the UK Biobank (UKB). Additionally, we studied the interaction between PRS and family history (FH) in the same population.

**Methods:** To calculate the PRS, we used a previously published panel of SNPs derived from the EUR population and applied it to the individuals of SAS ancestry from the UKB study. We applied the PRS using summary statistics from genome-wide association studies (GWAS) for cardiometabolic and lifestyle diseases such as coronary artery disease (CAD), obesity, and type 2 diabetes (T2D). Each PRS was adjusted according to an individual’s predicted genetic ancestry to derive an adjusted PRS (aPRS). We calculated the percentiles based on aPRS and divided them according to the percentiles into three categories: low, intermediate, and high. Considering the intermediate-aPRS percentile as a reference, we compared the low and high aPRS categories and generated the odds ratio (OR) estimates.

**Results:** The risk of developing severe obesity for individuals of SAS ancestry was almost threefold higher for individuals with high aPRS than for those with intermediate aPRS, with an OR of 3.67 (95% CI = 2.47-5.48, P < 0.01). While the risk of severe obesity was lower in the low-aPRS group (OR = 0.19, CI = 0.05–0.52, P < 0.01). Comparable results were found in the EUR data, where the low-PRS group had an OR of 0.26 (95% CI= 0.24-0.3, P < 0.01) and the high-PRS group had an OR of 3.2 (95% CI = 3.1-3.3, P < 0.01). We observed similar results for CAD and T2D. Further, we show that SAS individuals with a familial history of CAD and T2D with high-aPRS exhibit further higher risk to these diseases, thereby implying a greater genetic predisposition to these conditions.

**Conclusion:** Our findings suggest that using CAD, obesity, and T2D GWAS summary statistics predominantly from the EUR population have sufficient power to identify SAS individuals with higher genetic risk. With future GWAS recruiting more SAS participants and tailoring the PRSs towards SAS ancestry, we believe that the predictive power of PRS would improve.

## Background

Several genome-wide association studies (GWAS) for more than 5000 traits in GWAS Catalog(1) have been conducted to date, and very few of the GWASs have had significant success translating into the clinical setting (2). Hence, it is a significant milestone to translate GWAS findings to clinical settings, particularly for traits with high heritability. One of the drawbacks of the GWAS findings is that the identified genome-wide significant SNPs do not have such a large effect size in most cases. However, a current approach of combining those SNPs to a single score known as a polygenic risk score (PRS) has become popular to enhance the accuracy of predicting individuals at risk and has thus shifted the focus of the genetic community towards the use of GWAS findings again(3). PRS can be a precious tool for risk stratification, particularly in identifying groups of people with extremely high or low genetic risk of developing a specific disease or trait. Moreover, based on our recent work and others, it has become clear that for certain traits high PRS along with rare disease-causing variants can further increase the individuals’ risk of developing a disease compared to carriers without a high PRS (4–7).

Identifying the risk SNPs using GWAS requires a considerable sample size as even most disease-related SNPs have relatively small effect sizes. Today, most of the larger GWASs are mainly conducted with individuals with European (EUR) ancestries. Although improving polygenic prediction in non-EUR populations are needed, studies on South Asian (SAS) population become the second most studied ancestry group after EURs. Other than that, all other ancestries constitute less than 5% of GWAS. This bias in GWAS results impedes the use of PRS on an absolute scale for several complex traits in non-EUR populations (8). Thus, despite ongoing efforts to increase global genetics research diversity, it seems that it will take still some time to attain sufficient GWAS sample sizes to identify population-specfic risk SNPs.

As mentioned earlier, PRS is a potent tool to identify the sub-populations at risk. However, this inability to use it across populations with different ancestries is an important research topic. This lack of portability of PRS is due to differences in linkage disequilibrium (LD), risk variants, effect sizes, and allele frequencies. Further, methods to genotype or impute the missing SNPs initially developed with EUR ancestry in mind can increase those differences (9).

Several studies were being performed to study the portability of EUR-derived PRSs into other ancestries and an SAS specific PRS has been developed for CAD using previously published GWAS statistics (10). However, the majority of them had limited success (11**–**13). The PRS derived from EUR performed poorly in African population (14) and similar results were observed in a Latino/Hispanic population for some traits (15). While EUR-derived PRSs showed similar results for quantitative traits like blood count and anthropometric features, it performed poorly for blood pressure traits (16). Others have shown a connection between PRS and genetic ancestry (11,17). In other words, the studies show that applying PRSs derived from the EUR population directly on other ancestries might not be ideal. However, few studies used an approach to developing an ancestry-adjusted PRS (aPRS) that is mainly derived from EUR and can be transferred to other ethnicities (18). For example, a study showed a compromised solution where they found a minimal decrease in the prediction power of the PRS in SAS compared to EUR (19). Recently, it has been shown that in breast cancer, the PRS derived from EURs with an ancestry correction performed well in the SAS (13).

Compared to other ethnicities, SAS are at an increased risk of developing complex diseases such as coronary artery disease (CAD), obesity, and type 2 diabetes (T2D) (20). However, it is still unclear to what extent populations of EUR and SAS ancestry share the same genetic underpinnings of such cardiometabolic/lifestyle traits. The interplay between PRS, and family history (FH) in predicting the risk of various diseases has been a topic of interest in recent years (5,21–23). Although previous studies have examined the independent effects of FH and PRS, there is a lack of systematic research on the relative contributions and overlap of these factors across different types of familial risk in SAS.

Here, we systematically assessed the portability of the aPRS derived from EUR ancestry for obesity, CAD, and T2D to the SAS population and the interplay of FH and PRS in the same individuals. Hence, we used a published list of SNPs derived from the PGS catalog(24), then generated the aPRS and applied it to the EUR and SAS samples from the UK Biobank (UKB).

## Methods

### Data Source

The UKB is a prospective study that collects data over a long period and recruits volunteers aged between 40 and 69, mostly from Scotland, Wales, and England, totaling over 500,000 individuals. All participants have provided written consent and collected data is available for research purposes. The UK Biobank Axiom Array was used to generate genotyping data, which included around 850,000 variants and the imputation of over 90 million variants (25).

### Study cohort

CAD and T2D diagnoses were based on self-reported illness codes and ICD-10 diagnosis codes, including hospitalization records. Diagnosis of obesity was based on body mass index (BMI), with individuals having a BMI > 30 were considered obese. The UKB conducted quality control for the genetic data, and processed files were used in downstream analysis. We analyzed individuals of EUR and SAS ancestry and samples with discordant genotypic versus reported sex, sex chromosome aneuploidy, and high heterozygosity or missing genotype rates were considered as outliers (coded as “YES” in the fields 22001, 22019, and 22027 respectively) and excluded from further analysis. The outliers are defined as those coded as “YES” in the fields 22019, 22027, and 22001 respectively. We included only individuals who are unrelated up to the second degree,and from each pair of related individuals, one member was randomly retained (kinship coefficient > 0.0884, according to the UKB).

### Polygenic risk score analysis

PRSs were calculated using panel of SNPs identified in the previous studies (3,26) and the effect sizes were downloaded from PGS catalog (21) using the ids PGS000027, PGS000013, PGS000014 for BMI, CAD and T2D respectively. PRSice-2 was used to generate the PRS, which account automatically for allele-flipping and removing ambiguous SNPs (27). Strand-ambiguous SNPs are the ones with A/T or C/G alleles. Since many GWAS studies do not report the strand assignments, it is a standard practice in PRS calculations to exclude ambiguous SNPs. Since we already obtained the list of SNPs for the PRS calculation, we utilized the *‘–no clumping’* and *‘–no regress’* parameters along with the other default parameters, to bypass the time-consuming steps of regression and clumping. PRS values were standardized using the mean and standard deviation (PRS).

### Adjustment of PRS

Based on an previously applied approach (5,18) to reduce the variation in the PRS distribution due to genetic ancestry, we calculated an adjusted PRS (aPRS). A linear regression model was fitted using the first four principal components (PC) of ancestry derived from UKB (PRS ∼ PC1 + PC2 + PC3 + PC4). A predicted PRS was calculated based on this model. Finally, the aPRS was calculated by subtracting the predicted PRS from the raw PRS and standardized using the mean and standard deviation.

### Statistical analysis

After generating the aPRS, the next step involved stratifying SAS individuals based on aPRS percentiles. We divided individuals by their aPRS percentiles into three groups: <20% (low), 20-80% (intermediate), or >80% (high). First, we used logistic regression to compare low and high aPRS categories to an intermediate reference group, adjusting for age, sex, and the first four ancestry PCs. Second, we calculated odds ratios to determine the combined effect of aPRS, family history (FH), and disease development, using the “Intermediate” aPRS samples without FH as the reference group and adjusting for covariates such as sex, age, and the first four ancestry PCs.

### Model performance

For assessing the performance of the different models, the area under the curve (AUC) was used. Using the R package *pROC*, estimates with 95% confidence intervals (CIs) and AUC were computed. We divided the data to (75%) training and (25%) testing datasets. Logistic regression was used on the training data set, and model prediction and AUC calculations were made using the testing data set.

### Survival analysis

To calculate the lifetime cumulative risk based on PRS strata and FH status, a Cox proportional hazard model was used. Age served as the time scale, indicating span between the age at diagnosis and the event for observed cases and the age at the most recent visit for censored controls. Adjusted survival curves were produced considering the aPRS, age, sex, FH, and the first four ancestry PCs. R packages *survival* and *survminer* were used to perform Cox proportional hazard models, and R 4.2.2 was used for all statistical calculations.

## Results

### UK biobank dataset description

We identified a total 24,156 CAD cases among individuals of EUR ancestry and 822 SAS cases, with a mean age of 61.51 and 58.71 years at recruitment, respectively. The remaining individuals were considered controls. For T2D, we identified 25,526 cases among EUR individuals and 1,718 cases among SAS individuals, with a mean age of 60.39 and 57.42 years, respectively. For obesity, we identified 8,463 EUR and 105 SAS cases, with a mean age of 55.73 and 53.6 years, in EUR and SAS respectively (Table1).

**Table 1:** Characteristics of the participants by CAD, T2D, and Obesity diagnosis. coronary artery disease (CAD), type 2 diabetes (T2D), European (EUR), South Asian (SAS)

|  | **CAD** |  |  |  | **T2D** |  |  |  | **Obesity** |  |  |  |
| --- | --- | --- | --- | --- | --- | --- | --- | --- | --- | --- | --- | --- |
| *Ethnicity* | **EUR** |  | **SAS** |  | **EUR** |  | **SAS** |  | **EUR** |  | **SAS** |  |
| *Diagnosis* | Cases | Controls | Cases | Controls | Cases | Controls | Cases | Controls | Cases | Controls | Cases | Controls |
| *Participants, N* | 24156 | 428610 | 822 | 7842 | 25526 | 427240 | 1718 | 6946 | 8463 | 442811 | 105 | 8358 |
| *Male, N (%)* | 18514<br>(76.64) | 188835<br>(44.06) | 690<br>(83.94) | 3959<br>(50.48) | 15756<br>(61.73) | 191593<br>(44.84) | 1055<br>(61.41) | 3594<br>(51.74) | 2770<br>(32.73) | 203841<br>(46.03) | 29<br>(27.62) | 4457<br>(63.33) |
| *Female, N (%)* | 5642<br>(23.36) | 239775<br>(55.94) | 132<br>(16.06) | 3883<br>(49.52) | 9770<br>(38.27) | 235647<br>(55.16) | 663<br>(38.59) | 3352<br>(48.26) | 5693<br>(67.27) | 238970<br>(53.97) | 76<br>(72.38) | 3901<br>(46.67) |
| *Age, mean (SD)* | 61.51<br>(6.19) | 56.53<br>(8.03) | 58.71<br>(7.69) | 53.05<br>(8.35) | 60.39<br>(6.77) | 56.58<br>(8.04) | 57.42<br>(7.79) | 52.64<br>(8.34) | 55.73<br>(7.64) | 56.81<br>(8.02) | 51.99<br>(8.11) | 53.6<br>(8.47) |
| *Family history of CAD, N (%)* | 14759<br>(61.1) | 188340<br>(43.94) | 457<br>(55.6) | 3193<br>(40.72) | 13131<br>(51.44) | 189968<br>(44.46) | 754<br>(43.89) | 2896<br>(41.69) | 4276<br>(50.53) | 198168<br>(44.75) | 50<br>(47.62) | 3520<br>(42.12) |
| *Family history of T2D, N (%)* | 5290<br>(21.9) | 90086<br>(21.02) | 431<br>(52.43) | 4144<br>(52.84) | 10120<br>(39.65) | 85256<br>(19.96) | 1113<br>(64.78) | 3462<br>(49.84) | 2737<br>(32.34) | 92324<br>(20.85) | 69<br>(65.71) | 4420<br>(52.88) |

In the SAS population, there were significantly more CAD cases with a positive FH of CAD (55.6%) than controls (40.72%) with OR 1.98 [1.70-2.31], P < 0.01. Comparing cases with a positive FH of T2D (64.78%) to controls with positive FH (49.84%), CAD were found to be significantly more common (OR = 2.09 [1.86-2.34], P < 0.01) in cases with a positive FH.

### Ancestry correction and PRS distribution within the UKBB cohort

When studying individuals of a particular ancestry, it is crucial to apply ancestry correction using principal components (PCs) derived from the reference population. This step ensures a normal distribution of both PRS and aPRS (aPRS_CAD, aPRS_T2D, and aPRS_BMI) as shown in Figure 1. However, when using only PRS without any ancestry correction, we observed a striking difference in the number of individuals assigned to high PRS (where high PRS was defined as an individual belonging to a PRS percentile >80%). Specifically, there were significant variations between ethnic groups (EUR and SAS). For example, 18.5% of EUR samples (83,955/452,766) had a high PRS, while almost all SAS samples (96.2%, 8,331/8,664) showed a high PRS. However, applying aPRS reduced this variation. For instance, 20% of EUR samples (90,627/452,766) and 19.2% of SAS samples (1,659/8,664) had a high aPRS, leading to a more comparable distribution of PRS across ethnic groups. Similar results have been observed for CAD and Obesity as well (Table 2). Our findings are in line with a previous study where they show that ancestry correction is crucial to place an individual in the correct aPRS percentile for disease risk prediction.

**Table 2:** Comparison of the distribution of (a) PRS (defined as PRS percentile >80%). Coronary artery disease (CAD), type 2 diabetes (T2D), European (EUR), South Asian (SAS)

|  | Ancestry correction | EUR samples with High PRS | SAS samples with High PRS |
| --- | --- | --- | --- |
| **T2D** | **PRS** | 83955 (18.5%) | 8331 (96.2%) |
|  | **adjusted PRS (aPRS)** | 90627 (20%) | 1659 (19.2%) |
| **CAD** | **PRS** | 89,438 (19.8%) | 2,848 (32.9%) |
|  | **adjusted PRS (aPRS)** | 90,440 (20%) | 1,846 (21.3%) |
| **Obesity** | **PRS** | 85853 (19%) | 6433 (74.3%) |
|  | **adjusted PRS (aPRS)** | 90794 (20.1%) | 1492 (17.2%) |

**Figure 1:**
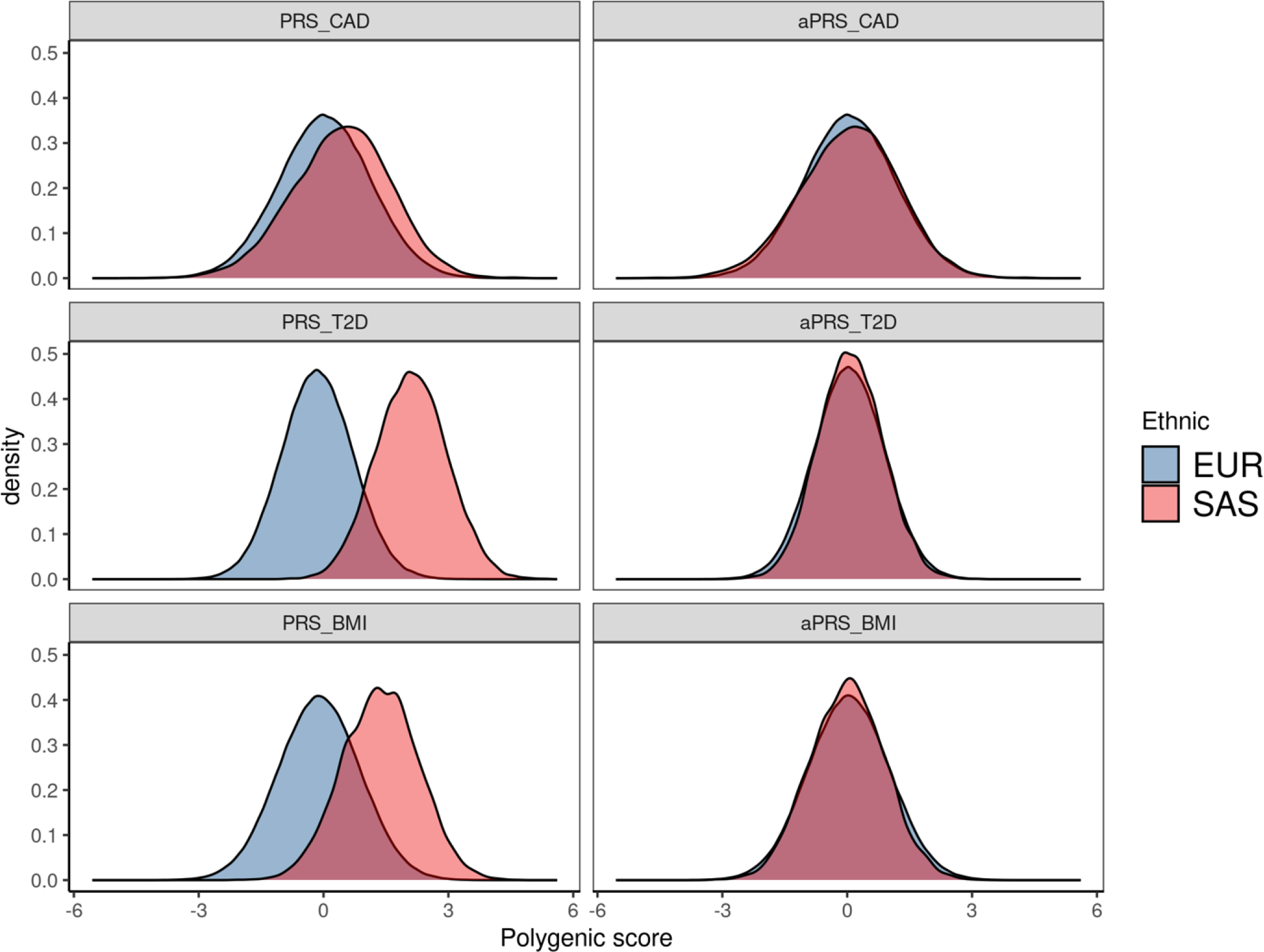
The distribution of PRSs before and after ancestry corrections across the various diseases. European (EUR), South Asian (SAS), coronary artery disease (CAD), type 2 diabetes (T2D), and adjusted polygenic risk scores (aPRS).

We next examined the performance of aPRS on SAS individuals. The aPRS were significantly higher in cases than in controls for all three phenotypes. The ORs of aPRS as a continuous variable were 1.53 (95% CI, 1.41-1.65, P < 0.01), 1.48 (95% CI, 1.37-1.60, P < 0.01), and 3.34 (95% CI, 2.59-4.34, P < 0.01) in SAS for T2D, CAD and obesity respectively. These ORs were comparable to the ones using the EUR PRS, which were 1.81 (95% CI, 1.78-1.84, P < 0.01), 1.67 (95% CI, 1.64-1.10, P < 0.01), and 2.62 (95% CI, 2.56-2.69, P < 0.01) for T2D, CAD and obesity respectively.

In the SAS population, aPRS improved the model discrimination over age, sex, and PCs (covariates). The AUCs for obesity, CAD and T2D obtained from the models that included PRS and covariates (0.79 (95% CI, 0.75-0.83), 0.79 (95% CI, 0.77-0.8), 0.69 (95% CI, 0.68-0.7)) were higher than those obtained from models based on only the covariates (0.67 (95% CI, 0.62-0.72), 0.76 (95% CI, 0.75-0.78), 0.67 (95% CI, 0.66-0.68)). The AUCs for obesity, CAD, and T2D were comparable to the EUR-based models including PRS and covariates: 0.74 (95% CI, 0.73-0.75), 0.78 (95% CI, 0.78-0.79), 0.7 (95% CI, 0.7-0.71) (Figure 2).

**Figure 2:**
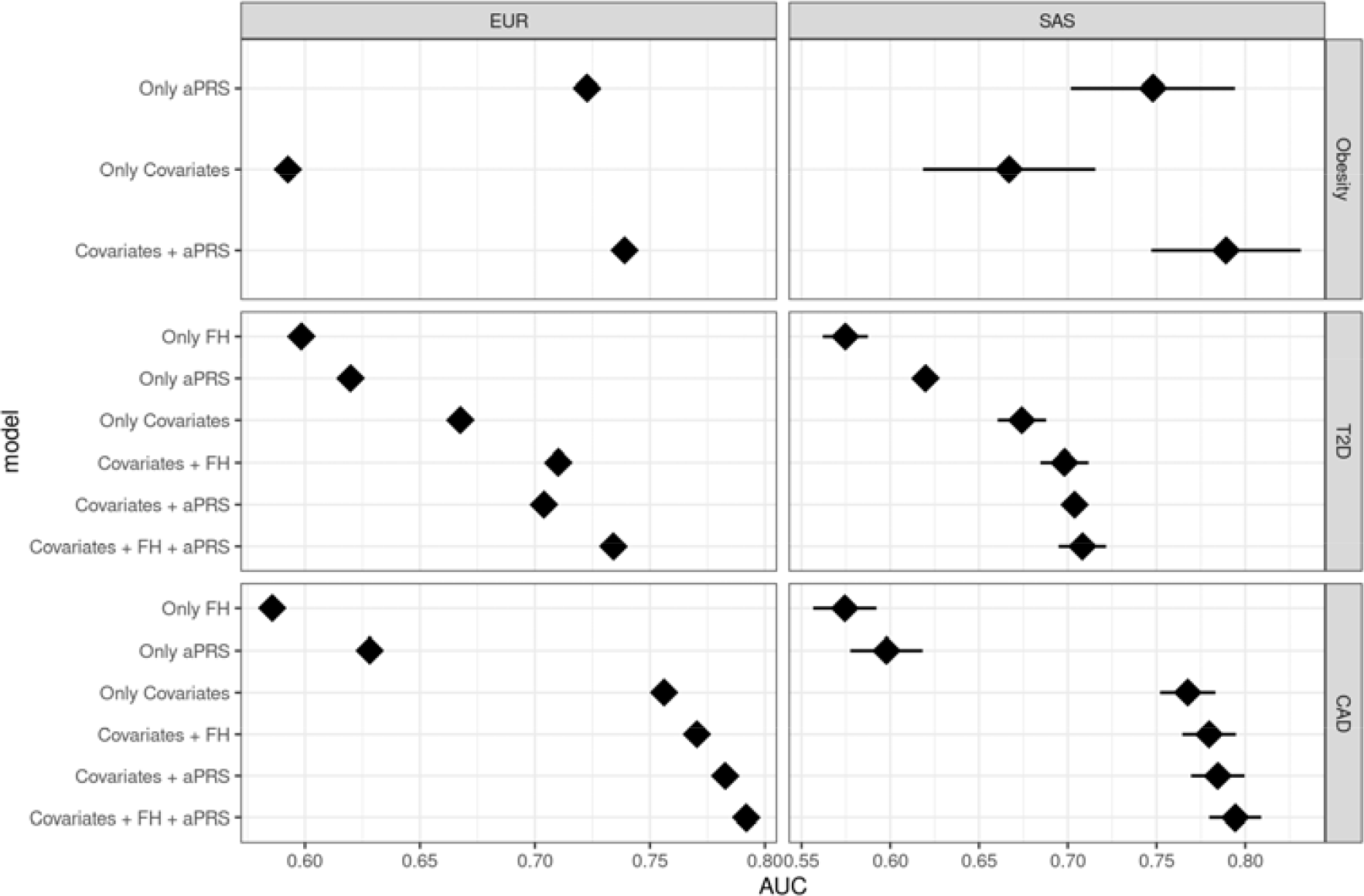
Comparison of models AUC including aPRS and covariates (age, sex, first four principal components). European (EUR), South Asian (SAS), coronary artery disease (CAD), type 2 diabetes (T2D), and adjusted polygenic risk scores (aPRS).

We stratified individuals based on their PRS or aPRS into three groups: low (< 20%), intermediate (20-80%), or high (> 80%) risk. In the SAS population, using the intermediate aPRS as reference, the CAD risk ranged from 0.56 (95% CI, 0.45–0.7) for individuals with low aPRS to 1.72 (95% CI, 1.44–2.05) for individuals with high PRS. The ORs for CAD based on PRS strata in SASs was comparable to those in EURs; the CAD risk ranged from 0.53 (95% CI, 0.51–0.56) for those with low aPRS to 2.06 (95% CI, 2.0–2.12) with high aPRS. The same trend was also seen for obesity and T2D. In SASs, people in the high obesity risk group had an OR of 3.67 (95% CI, 2.47–5.48), comparable to the one for high EUR risk of 3.2 (95% CI, 3.05-3.33). T2D risk was calculated in SASs in the high group as 1.55 (95% CI, 1.36–1.77) and as 1.87 (95% CI, 1.81-1.92) in EURs (Figure 3).

**Figure 3:**
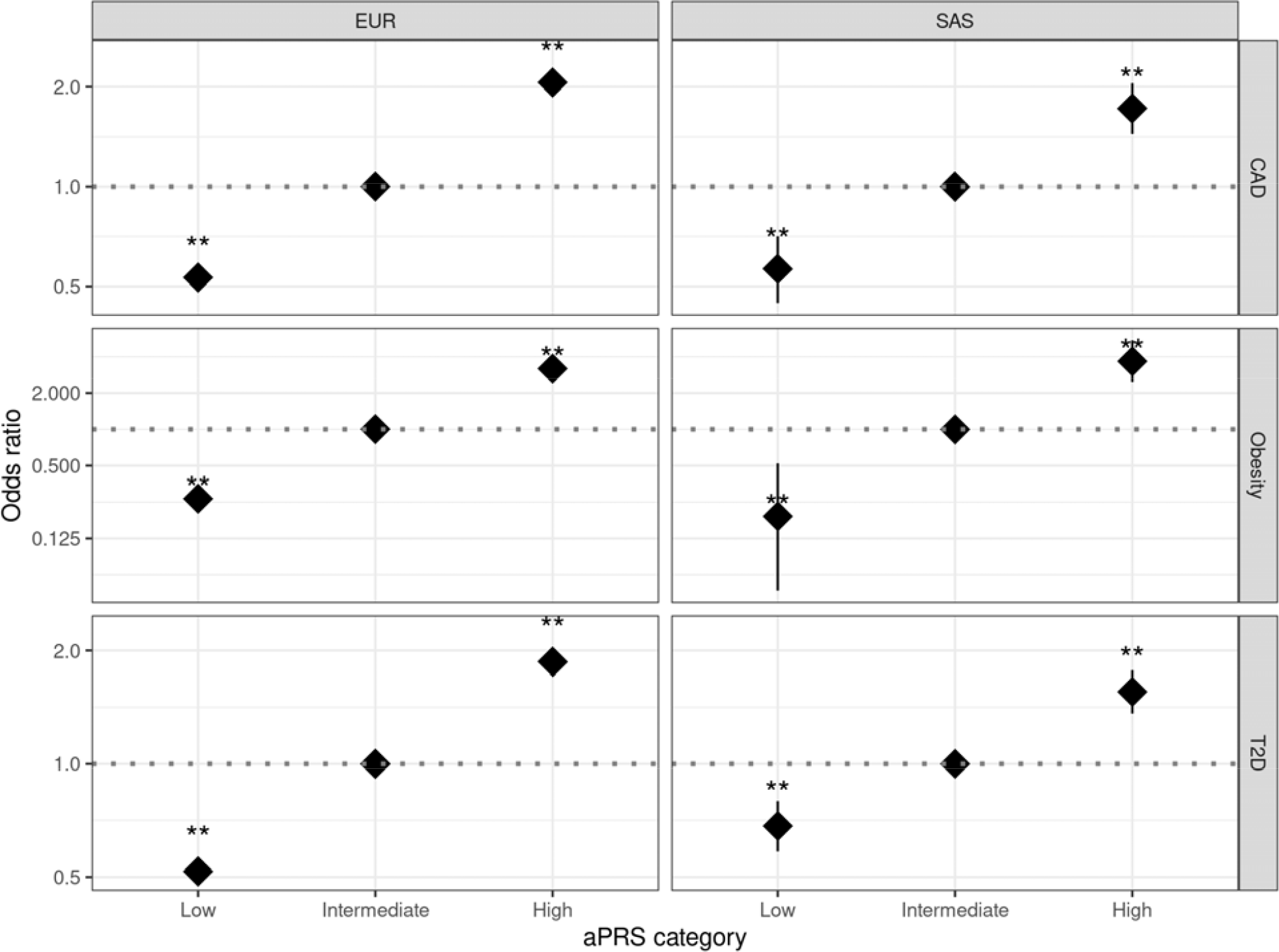
Odds ratio for CAD, T2D, and Obesity based on aPRS strata. European (EUR), South Asian (SAS), coronary artery disease (CAD), type 2 diabetes (T2D), and adjusted polygenic risk scores (aPRS). If a p-value is less than 0.01, it is flagged with two stars (**).

Individuals with a positive FH and high aPRS showed a higher risk of developing CAD compared to those with no FH and intermediate PRS (Figure 4). This trend was comparable in both SAS and EUR populations. In SAS, those with both positive FH and high aPRS had a five-fold increased OR of developing CAD compared to those with low aPRS and no FH, 3.44 (95% CI, 2.7-4.4). This was comparable to CAD ORs for EURs, ranging from 0.6 (CI=0.52-0.6) with no FH and low PRS to 3.85 (95% CI, 3.73-4.01) with FH and high PRS. No significant interaction was observed between FH status and PRS in both EUR and SAS individuals (p=0.12, and p=0.11, respectively). Notably, in SAS and EUR, individuals with negative FH and high aPRS had comparable risks of developing CAD as those with positive FH and intermediate aPRS (2-fold risk). However, individuals with low aPRS, even with a positive FH, had a CAD risk comparable to the reference group. The same trend was also shown in T2D.

**Figure 4:**
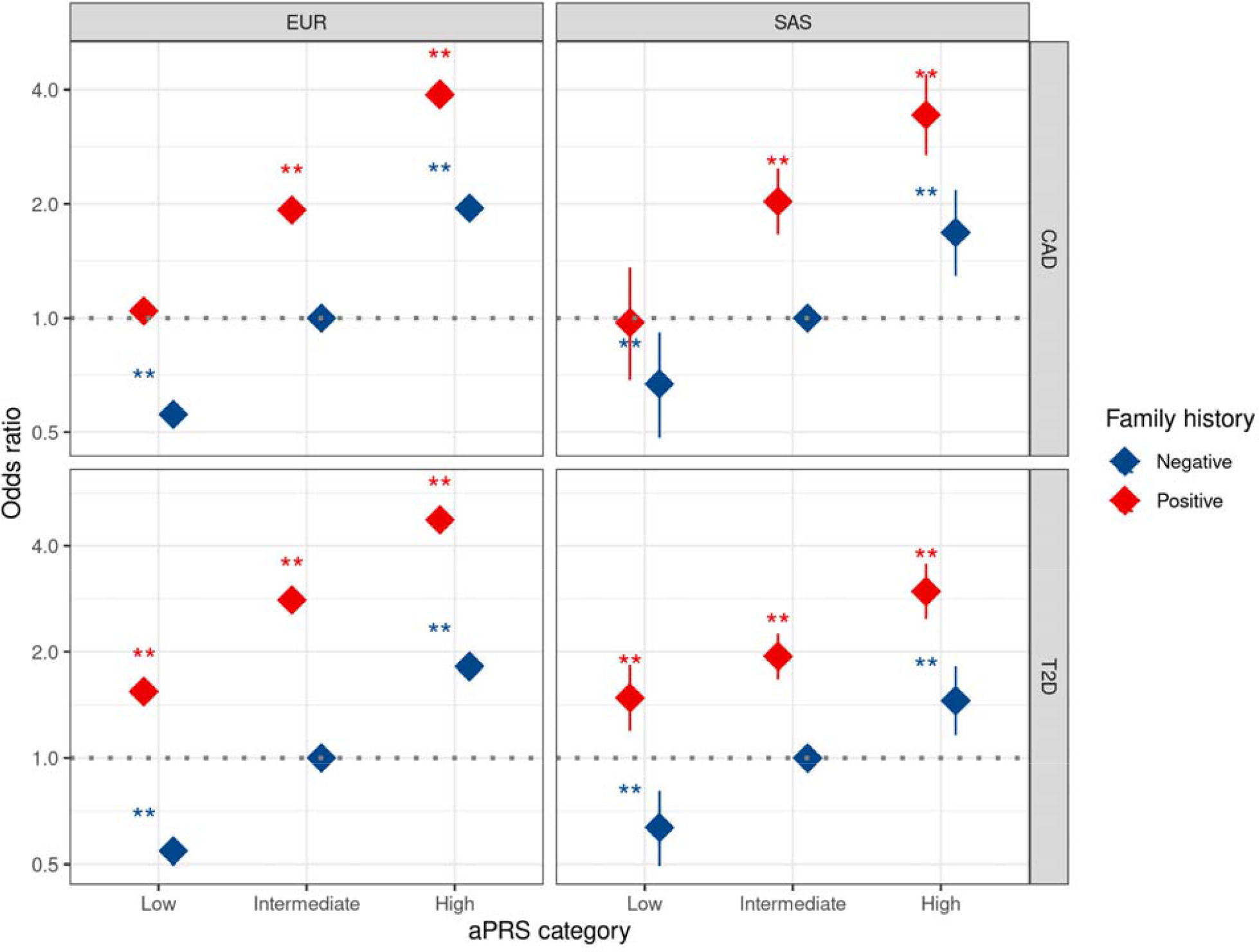
Odds ratio for CAD, T2D, and obesity based on aPRS strata and FH status. European (EUR), South Asian (SAS), coronary artery disease (CAD), type 2 diabetes (T2D), family history (FH) and adjusted polygenic risk scores (aPRS). If a p-value is less than 0.01, it is flagged with two stars (**).

The cumulative CAD incidence among SAS with positive FH increases from 46% with low aPRS to 75% with high aPRS by age 70 (Figure 5). Notably, SAS individuals with an intermediate aPRS and a positive FH had a cumulative CAD incidence (65%) comparable to those with a high aPRS and a negative FH (63%). The cumulative incidence of T2D among SAS individuals ranges from 58% with a negative FH and low aPRS to 95% with a positive FH and high aPRS. Cumulative incidence of T2D among individuals with high-aPRS of SAS ancestry (95%) were higher than EUR individuals (70%) in the corresponding PRS groups. A similar trend was observed in CAD.

**Figure 5:**
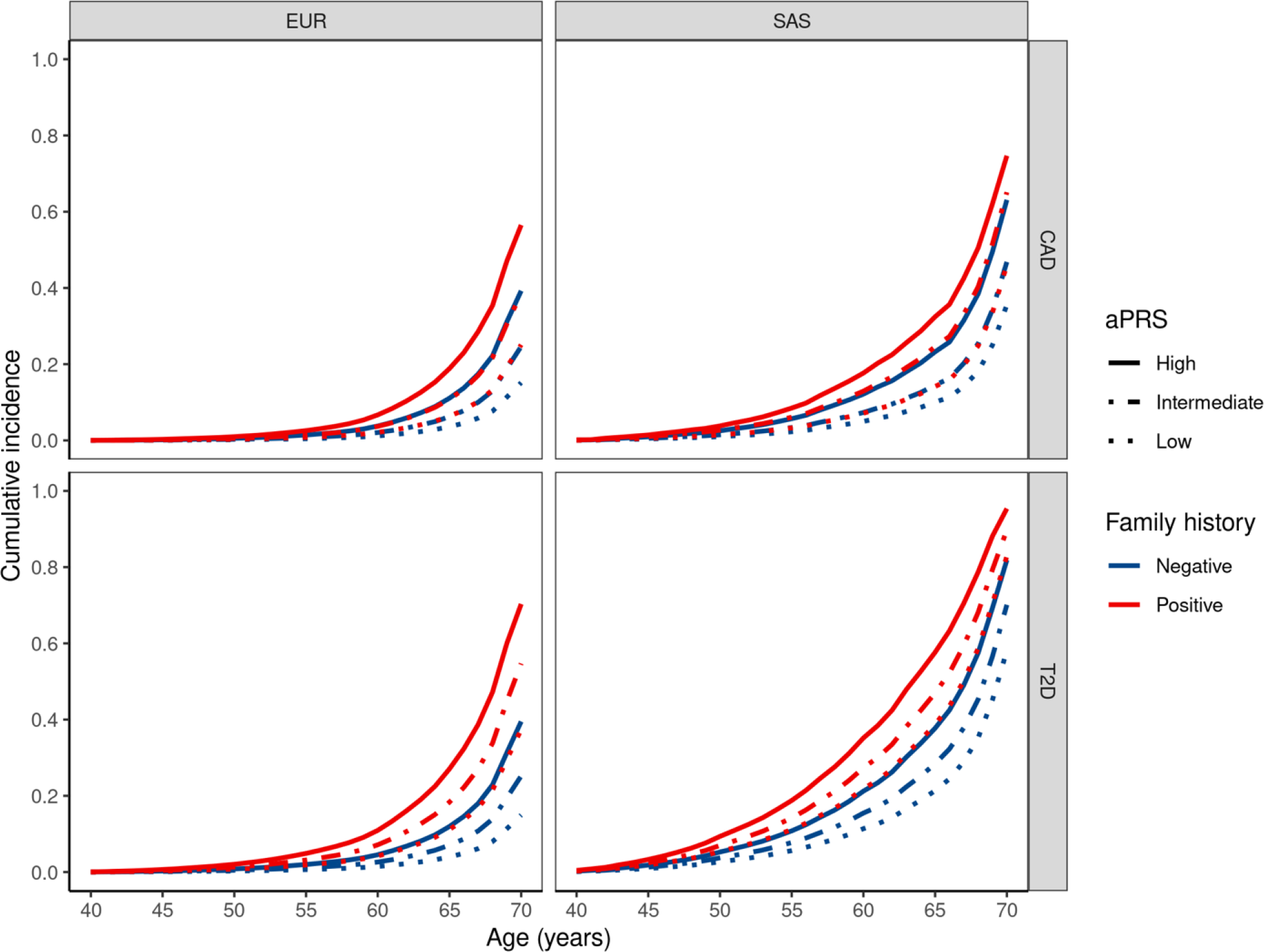
Cumulative incidence of CAD, T2D, and obesity based on aPRS strata and FH status. European (EUR), South Asian (SAS), coronary artery disease (CAD), type 2 diabetes (T2D), family history (FH) and adjusted polygenic risk scores (aPRS).

## Discussion

Extending the previous studies, we aimed to investigate the transferability of EUR-derived PRSs to the SAS population and explore the relationship between PRS and FH in contributing to the burden of CAD, T2D and obesity. The findings of this study using UK Biobank data demonstrate that an aPRS based on a large-scale GWAS of cardiometabolic diseases in EUR ancestry can identify individuals with high risk of disease predisposition in the SAS population in a comparable manner to those of EUR ancestry. Additionally, the aPRS stratifies SAS individuals with and without positive FH for both T2D and CAD, and a low aPRS compensates for the effect of FH, while a high PRS being linked to a considerably elevated risk. Among high aPRS individuals with positive FH, we noticed an increased cumulative incidence in individuals of SAS ancestry compared to EUR individuals stratified by PRS (Figure 5).

It has been shown that the UKB is a valuable resource for evaluating the utility of PRS, as it provides both phenotypic and genotypic data (28). While the majority of UKB participants have EUR ancestry, the dataset involves more than 20,000 participants of self-reported non-EUR.

However, a major challenge with using PRS in clinical settings is that the distribution of genetic variants can vary widely among different ethnic populations (8). This can result in inaccurate predictions of disease risk and hinder the validation of PRS in diverse populations (see Figure 1). The observed dissimilarity between the distributions for EURs and SASs highlights the need of adjusting for the correct ancestral background to accurately assign an individual to their respective percentile within the reference distribution.

To address this issue, we have used a technique called population structure adjustment (19), which involves accounting for the genetic differences between different populations when calculating PRS. By adjusting for population structure, we minimized the impact of genetic variability on the accuracy of PRS predictions and facilitate the validation of PRS in diverse populations.

The generalizability of the study’s findings is subject to limitations stemming from several factors. The study participants were recruited exclusively within the UK, including individuals of EUR and SAS ancestry. Thus, healthcare access and non-genetic risk factors may be more comparable among these ethnic groups as they would be expected using two cohorts recruited in EUR and SAS separately. Nevertheless, it is important to acknowledge that socioeconomic determinants, lifestyle choices, and health disparities may differ across various ethnic groups even living in the same region. Although certain risk variants are likely specific to certain populations, the findings indicating similar performance of the PRS across ancestry groups suggest that non-EUR groups, including SAS, may share some of the identified risk variants found in EUR-based GWAS for cardiometabolic disorders.

The findings of our study reveal that a higher PRS was found to be associated with an increase in obesity, T2D and CAD cases among individuals of SAS ancestry. However, the performance of the EUR-based PRSs was observed to be inadequate in the African (AFR) population, suggesting the existence of ancestry-specific differences (29). Hence, PRSs should be evaluated carefully by ancestry groups to assess their transferability across ancestries and diseases. Whenever possible PRS should be constructed based on GWAS based on the same ancestry group(30).

PRS based on EUR GWAS may not be ideal for all diseases in non-EUR groups but can aid in risk assessments for some diseases(31). Various strategies, such as deferring implementation until ancestry-specific GWAS are available or modifying current PRS with clear limitations for specific individuals or groups, can be used to evaluate PRS usefulness. Methodological advancements utilizing local ancestry or GWAS statistics from diverse populations will enhance the performance of PRS across different ancestry groups. In the meanwhile, we validated the efficacy of the latter strategy for risk categorization in SAS individuals using and adjusted PRS from the EUR population.

The increasing availability of data from larger and more diverse populations, coupled with technological advancements, has spurred interest in the clinical adoption of PRS. Recent research has demonstrated that combining clinical risk scores with PRS can help identify more people who are at risk of developing T2D, especially in SAS populations. Our study provides a potential model for laboratories and health systems seeking to utilize a EUR-derived PRS in SAS populations. Additionally, our study contributes to literature that supports the use of PRS and FH as complementary measures in assessing inherited disease susceptibility for T2D and CAD (5).

## Conclusion

Taken together, our study provides evidence supporting the transferability of EUR-derived PRSs to SAS populations for identifying individuals at high risk of T2D, obesity and CAD. Our findings emphasize the importance of considering both polygenic risk and family history in assessing disease risk in clinical practice, which can improve risk prediction and inform personalized prevention and management strategies for these common non-communicable diseases. To assess the clinical utility and cost-effectiveness of implementing these measures in diverse populations, further research is needed.

## Data Availability

Genome-wide genotyping data, exome-sequencing data, and phenotypic data from the UK Biobank are available upon successful project application (http://www.ukbiobank.ac.uk/about-biobank-uk/). Restrictions apply to the availability of these data, which were used under license for the current study (Project ID: 52446).

## Declarations

### Ethics approval and consent to participate

Not Applicable

### Consent for publication

Not Applicable

### Authors’ contributions

EH, and DRB performed the statistical analysis and the bioinformatics. EH, PM and DRB conceived and designed the study. EH, PM, and DRB drafted the initial manuscript. EH, PK, CM, PM, and DRB performed the critical expert revision. PK, CM, PM, and DRB supervised the study. All authors read and approved the final manuscript.

## Acknowledgements

UK Biobank analyses were conducted via application 52446 using a protocol approved by the Partners HealthCare Institutional Review Board. EH, DRB and PM were supported by the FNR INTER INTER/DFG/21/16394868.

